# Evaluation of Prognostic Utility of Tumor Mutation Burden for NSCLC response to Immune Checkpoint Blockade Therapy: A Single Institute Study

**DOI:** 10.1101/19013433

**Authors:** Jean R. Clemenceau, Sung Hak Lee, Peter Bazeley, Alex Millinovich, Jian Jin, Nathan Pennell, Davendra Sohal, Tae Hyun Hwang

## Abstract

**IMPORTANCE:** The role of Tumor Mutation Burden (TMB) as a prognostic and/or predictive biomarker for Immune Checkpoint Blockade (ICB) therapy in a real-world clinical setting is still unclear.

**OBJECTIVE:** To assess whether TMB status provided by a clinically and commercially available tumor genomic profiling (TGP) assay is associated with overall survival of Non-Small Cell Lung Cancer (NSCLC) patients treated with ICB from a single institute.

**DESIGN, SETTING, AND PARTICIPANTS:** Outcomes and genetic testing data were collected for 188 NSCLC patients treated within the Cleveland Clinic system between August 2012 and July 2017.

**MAIN OUTCOMES AND MEASURES:** Overall survival (OS) from time receiving ICB therapy.

**RESULTS:** Among 188 patients with NSCLC (median age, 62 years; 49.5% female), 86 (45.7%) received ICB therapy. Patients were grouped into three categories based on the status of TMB (in mutations/Mb): high (>= 20/Mb), intermediate (>=5 to <= 20/Mb), and low (<5/Mb). In patients treated with ICB, TMB high status was not significantly associated with improved OS from therapy initiation (HR: 0.90 [95% CI, 0.52-2.49, P>0.8], median OS difference: 9.7 months).

**CONCLUSIONS AND RELEVANCE:** Among patients with NSCLC from a single institute in a longitudinal database of clinical data including TGP results, exploratory analyses do not show statistical significance for the prognostic utility of TMB. These findings indicate that there is a need for more prospective data on the use of TMB status as a guide for ICB therapy in a routine care setting.

## Introduction

Tumor Mutation Burden (TMB), defined as the number of somatic mutations per megabase of DNA using whole-exome sequencing (WEX) or targeted genomic profiling (TGP), has been found to associate with Immune Checkpoint Blockade (ICB) response in advanced cancers including non-small cell lung cancer (NSCLC).^1^ In recent years, the FDA has approved genomic profiling tests for guiding cancer management, including Foundation Medicine’s Foundation One (FO) gene panel assay^2^, which includes results on TMB levels. Although, there is an increasing utilization of TGP such as the FO assay in standard clinical care, there are few observational investigations of the prognostic and/or predictive utility of TMB status reported on the clinically and commercially available TGP for ICB response in NSCLC patients in the real-world setting,^3^ and they still show inconsistencies with clinical trial data. Here we performed a retrospective analysis of the role of TMB classification reported in a clinical TGP assay in predicting differences in outcomes among NSCLC patients undergoing ICB therapy, using longitudinal data from a single hospital system within the United States.

## Methods

### Study Design

With approval from the Cleveland Clinic (CC) Institutional Review Board, demographic and clinical outcome data were collected on adult patients diagnosed with NSCLC who were treated at any CC site within the United States, and who received genetic testing from a FO assay between August 2012 and July 2017. FO test results are routinely reviewed by the CC Molecular Tumor Board for guidance on clinical care. Patient data was collected from the CC Electronic Health Record (EHR) system, and FO report data was collected from Foundation Medicine through their clinical data portal. All data was formatted and collected into a secure MySQL database locally stored within the CC Lerner Research Institute.

Patients were included if they were 18 years old or older at the time of the FO report date, and whose FO results included a pathologist’s diagnosis of NSCLC, defined as any of: large cell, squamous cell, or sarcomatoid carcinoma of the lung, or lung adenocarcinoma. From these initial inclusion criteria, additional filters were applied to maximize the quality of our analysis. Namely, we included only patients with no missing report date, biopsy date, or date of first diagnosis, for whom cancer therapy records including ICB were available, and whose endpoint (date of death or last follow up) was recorded after their report date. If patients had multiple FO reports, only the earliest report was considered for all analyses. Further, we limited TMB status criteria to somatic and likely somatic genomic alterations (GA), as determined by Foundation Medicine.

We calculated Overall Survival (OS) as the number of months after a patient’s FO report date until death of any cause or the date of last follow-up. For OS after immunotherapy, we considered the number of months after the earliest Immune Checkpoint Blockade (ICB) therapy order date. Metastasis status was determined as any diagnosis of metastases or secondary cancer occurring after the initial NSCLC diagnosis and before the endpoint. Treatment Naivety was determined by the number of cancer therapies the patient received before the FO report date. If more than one therapy was given, we considered the patient’s cancer as “refractory”. A patient was considered EGFR mutant positive if the FO report listed at least 1 GA in the EGFR gene.

Tumor Mutation Burden (TMB) was calculated as the total number of short genomic variants called by FM divided by the total number of non-overlapping megabases sequenced in the assay. FO reports before August 2014 comprised 0.91 Mb of targeted sequence, and reports between this date and September 2018 examined 1.25Mb. TMB was then categorized as TMB-Low if the value was at or below 5, TMB-Intermediate if between 5 and 20, and TMB-High if at or above 20 mutations per megabase.

### Statistical Analysis

The significance of association between categorical characteristics of the patients and TMB status was established using Pearson’s Chi-squared test. Differences in the distribution of age across the 3 TMB groups was assessed with a Kruskal–Wallis test. Kaplan-Meyer (KM) survival curves were plotted for the 3 TMB groups and compared using the Log-Rank test. To analyze OS post-ICB, subjects were dropped if they had an unknown ICB order date or OS less than 1 month.. Univariate Cox Proportional Hazards Regression (CoxPH) models were fit using TMB-Low as the reference group. Additionally, a multivariable CoxPH model was fit on OS for TMB status while adjusting for age at report date (age), sex, race, ethnicity, metastasis status, primary cancer cell type, EGFR mutation status, EGFR targeted therapy history, ICB therapy history, treatment naivety, and number of cancer therapies received before ICB treatment. These covariates were included to account for any confounding due to socioeconomic healthcare disparities,^4–6^ a mutation profile known to respond to other therapies, and evidence of resistance to therapies. Survival analysis was performed on the complete NSCLC dataset and the subset of patients who received ICB therapy at any point in their care at CC. These analyses were repeated on the collection of all CC patients who received ICB during the same timeframe.

All statistical analyses were 2 sided and evaluated using a Type I Error probability of α=0.05. The tests were performed and visualized with R (v3.3 R Foundation for Statistical Computing) using the following packages from CRAN: *stats, survival, survminer, and ggplot2*.

## Results

### Patient Characteristics

A total of 188 NSCLC patients were included in the study, of which 86 had undergone ICB therapy. A total of 78 NSCLC patients with ICB conserved all the required fields for further analysis. The majority of patients (130, 69.15%) were classified as TMBI, as well as 38 (20.21%) and 20 (10.64%) classified as TMB-High and TMB-Low, respectively. Demographically, 93 (49.47) of the patients were female, 153 (81.4%) where white, 177 (94.15%) were non-Hispanic, and the median age was 62 (IQR: 55-71; range: 25-89). The baseline characteristics are described in Table 1. The majority of these characteristics did not show significant differences among TMB categories, except for the association between EGFR mutation status (p = 0.03).

**Table 1.** Characteristics of all NSCLC patients in the study according to TMB status.

| Characteristic | TMB-High<br>(n=38) | TMB-Intermediate<br>(n=130) | TMB-Low<br>(n=20) | Total (n=188) | P Value |
| --- | --- | --- | --- | --- | --- |
| **Sex** |  |  |  |  | 0.75 |
| Female | 20 (52.63%) | 62 (47.69%) | 11 (55.00%) | 93 (49.47%) |  |
| Male | 18 (47.37%) | 68 (52.31%) | 9 (45.00%) | 95 (50.53%) |  |
| **Race** |  |  |  |  | 0.07 |
| Caucasian | 27 (71.1%) | 106 (81.5%) | 20 (100.0%) | 153 (81.4%) |  |
| Black | 9 (23.7%) | 16 (12.3%) | 0 (0.0%) | 25 (13.3%) |  |
| Other | 2 (5.26%) | 8 (6.15%) | 0 (0.00%) | 10 (5.32%) |  |
| **Ethnicity** |  |  |  |  | 0.92 |
| Not Hispanic or Latino | 36 (94.74%) | 122 (93.85%) | 19 (95.00%) | 177 (94.15%) |  |
| Hispanic American | 1 (2.63%) | 6 (4.62%) | 1 (5.00%) | 8 (4.26%) |  |
| Unknown Ethnic Group | 1 (2.63%) | 2 (1.54%) | 0 (0.00%) | 3 (1.60%) |  |
| **Age** | 66.5 (57-75)<br>[43-87] | 62.5 (55-71)<br>[25-89] | 59 (55-68.75)<br>[37-79] | 62 (55-71)<br>[25-89] | 0.36 |
| **Metastasis Status** |  |  |  |  | 0.17 |
| No Known Mets | 3 (7.9%) | 16 (12.3%) | 5 (25.0%) | 24 (12.8%) |  |
| Metastasis | 35 (92.1%) | 114 (87.7%) | 15 (75.0%) | 164 (87.2%) |  |
| **EGFR Status** |  |  |  |  | 0.03* |
| Wild Type | 36 (94.7%) | 108 (83.1%) | 20 (100.0%) | 164 (87.2%) |  |
| EGFR mutant | 2 (5.3%) | 22 (16.9%) | 0 (0.0%) | 24 (12.8%) |  |
| **EGFR Therapy** |  |  |  |  | 0.60 |
| Received EGFR Tx | 7 (18.4%) | 29 (22.3%) | 6 (30.0%) | 42 (22.3%) |  |
| No EGFR Tx | 31 (81.6%) | 101 (77.7%) | 14 (70.0%) | 146 (77.7%) |  |
| **ICB Therapy** |  |  |  |  | 0.11 |
| No ICB | 16 (42.1%) | 72 (55.4%) | 14 (70.0%) | 102 (54.3%) |  |
| Received ICB | 22 (57.9%) | 58 (44.6%) | 6 (30.0%) | 86 (45.7%) |  |
| **Tx Naivety** |  |  |  |  | 0.33 |
| Tx Naïve | 8 (21.1%) | 16 (12.3%) | 2 (10.0%) | 26 (13.8%) |  |
| Refractory | 30 (78.9%) | 114 (87.7%) | 18 (90.0%) | 162 (86.2%) |  |
| **Cancer Diagnosis** |  |  |  |  | 0.90 |
| Lung Adenocarcinoma | 28 (73.68%) | 98 (75.38%) | 14 (70.00%) | 140 (74.47%) |  |
| Large or NOS Cell Lung Carcinoma | 7 (18.42%) | 17 (13.08%) | 4 (20.00%) | 28 (14.89%) |  |
| Lung Squamous Cell Carcinoma | 2 (5.26%) | 13 (10.00%) | 2 (10.00%) | 17 (9.04%) |  |
| Lung Adenosquamous Carcinoma | 1 (2.63%) | 1 (0.77%) | 0 (0.00%) | 2 (1.06%) |  |
| Lung Sarcomatoid Carcinoma | 0 (0.000%) | 1 (0.769%) | 0 (0.000%) | 1 (0.532%) |  |

### Survival Analysis

Survival analysis of the complete NSCLC cohort with respect to TMB status indicated no statistically significant relationship. As observed in Figure 1a, survival curves for all groups follow a similar trend with a Log-Rank p=0.87 and a median survival difference of 1.71 months in TMB-Low vs TMB-Intermediate and 2.95 months in TMB-Low vs TMB-High. Univariate CoxPH analysis indicated non-significant worsened outcomes for TMB-Low vs TMBI, with a HR=1.20 (CI95: 0.60-2.37), and for TMB-Low vs TMB-High, with HR=1.15 (CI95: 0.54-2.47). Multivariate analysis (Figure 1b) replicated this non-significant decrease in OS for TMB-Intermediate HR=1.61 (CI95: 0.73-3.54) and TMB-Low HR=2.27 (CI95: 0.96-5.37). There were, however, significant OS improvements for Hispanic patients (HR: 3.07, CI95: 1.12-8.42) and those of unknown ethnic group (HR: 5.27, CI95: 1.06-26.26), patients diagnosed with NSCLC-NOS (HR: 2.99, CI95: 1.56-5.74), those with history of ICB therapy (HR: 2.69, CI95: 1.39-5.20), and those with refractory cancers (HR: 3.22, CI95: 1.47-7.05).

**Figure 1a.**
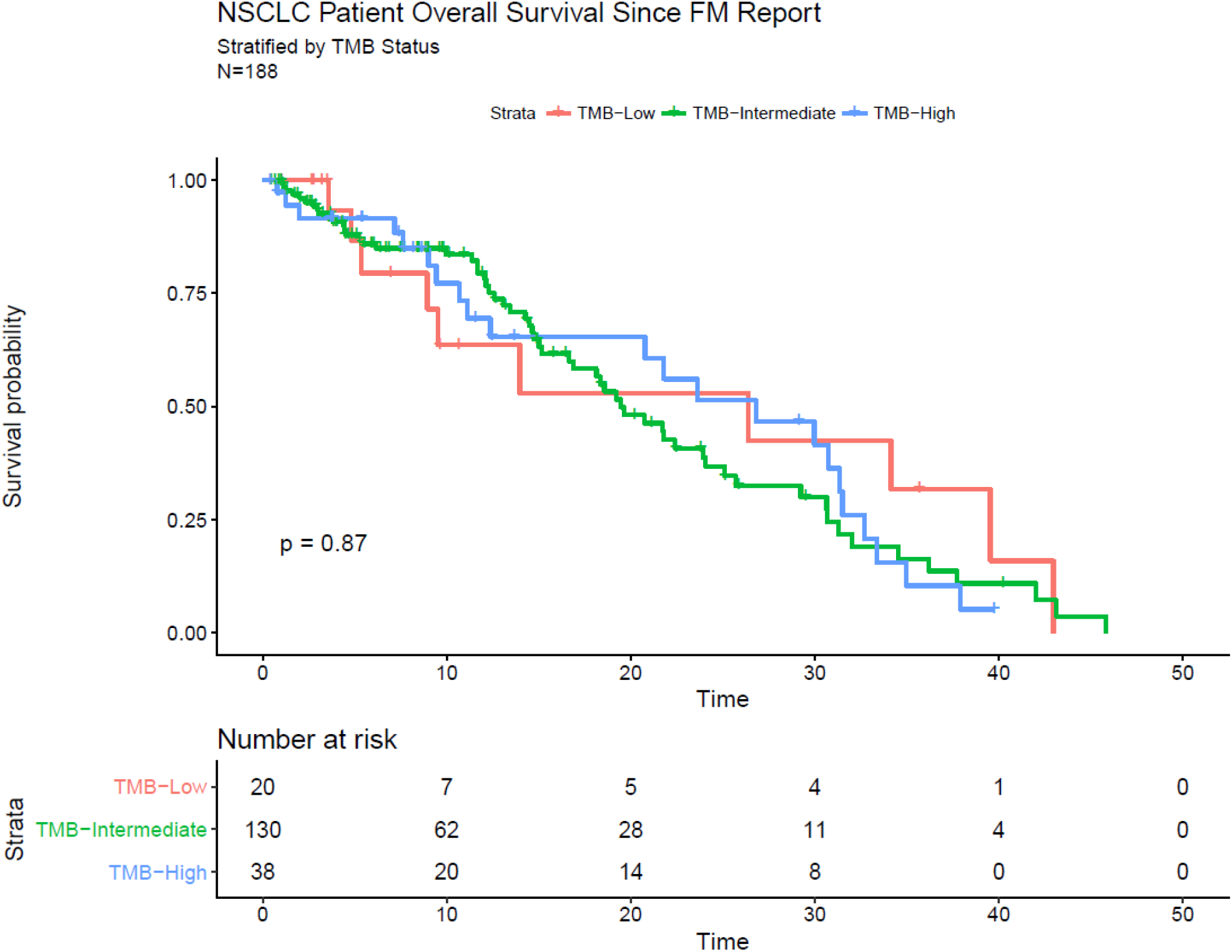
Overall Survival of NSCLC Patients according to TMB status.

**Figure 1b.**
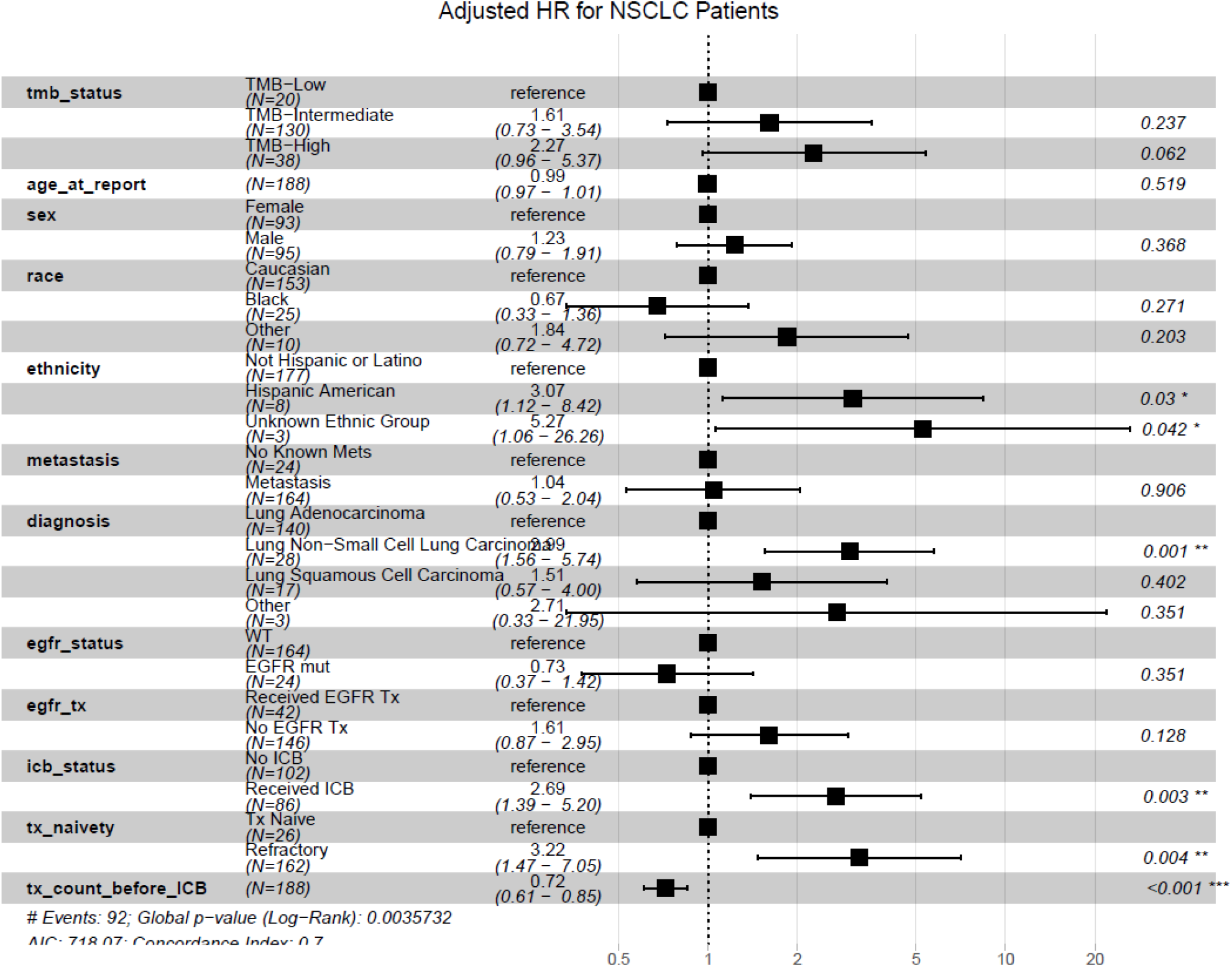
Hazard Ratios for TMB Status in NSCLC Patients Adjusted for Covariates.

Of the 86 patients who underwent ICB therapy, 8 patients had either an unknown ICB order date or had an OS less than 1 month. The median OS difference for TMB-High patients was 9.7 months less than TMB-Low patients, while TMBI patients had a median OS 11.48 months. Although these are considerable differences in median survival, Figure 2a shows that the rest of the distributions are fairly close and their differences are not statistically significant (p=0.55). Univariate CoxPH models also indicated no significant difference in outcomes compared to TMB-Low: HR=1.33 (CI95: 0.46-3.87) for TMBI, and HR=0.90 (CI95: 0.27-2.97) for TMB-High. The multivariate CoxPH (Fig 2b) showed non-significant worse outcomes for both TMBI (HR: 0.63, CI95: 0.18-2.19) and TMB-High (HR: 0.85, CI95: 0.20-3.58). On the other hand, males had significantly better HR than females (HR: 2.80, CI95: 1.24-6.32), as well as non-white or black patients (HR: 7.35, CI95: 2.07-26.09), while patients with diagnoses other than Lung Adenocarcinoma and NSCLC-NOS had worse outcomes (HR: 0.13, CI95: 0.02-0.78).

**Figure 2a.**
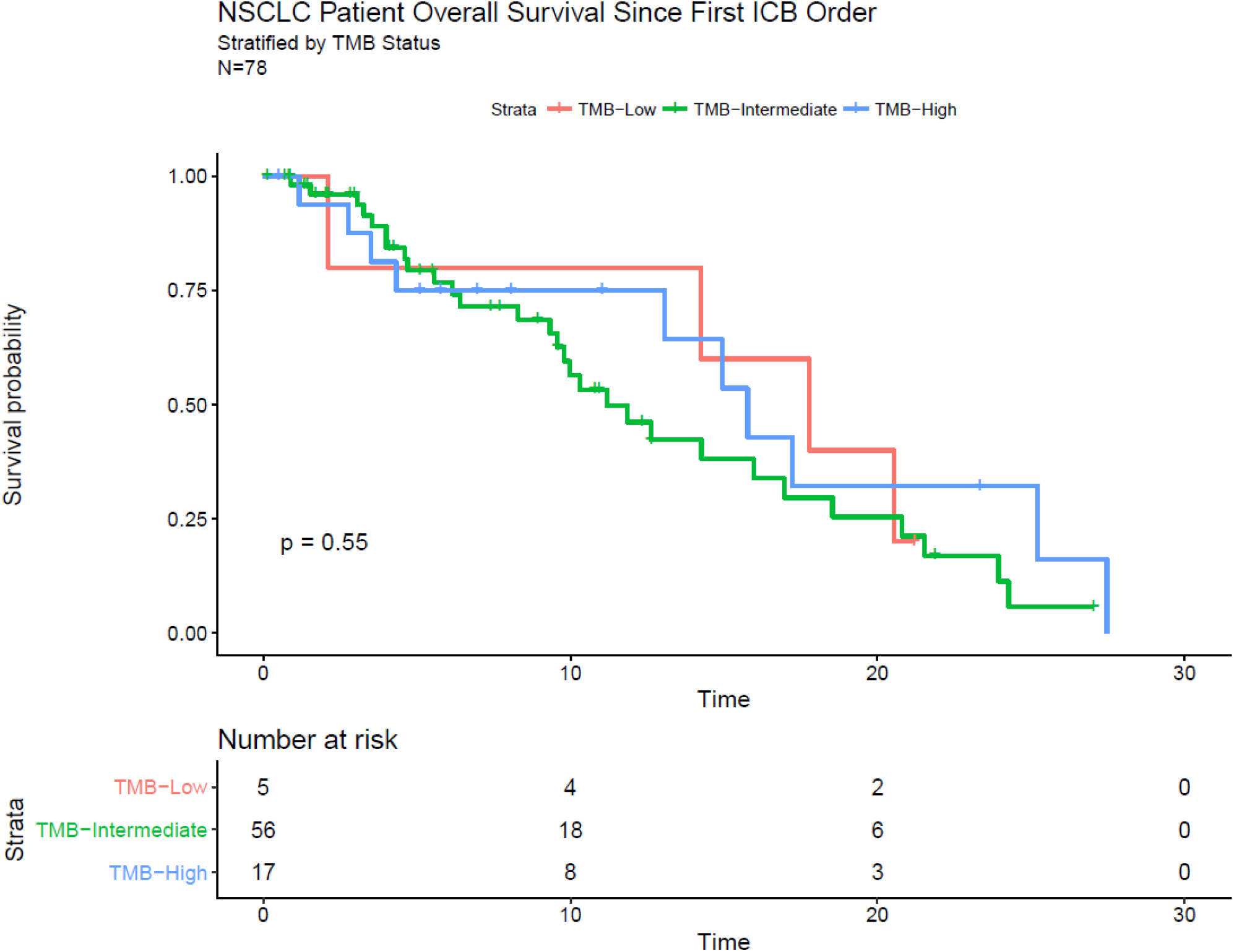
Overall Survival of NSCLC Patients with History of Immunotherapy According to TMB status.

**Figure 2b.**
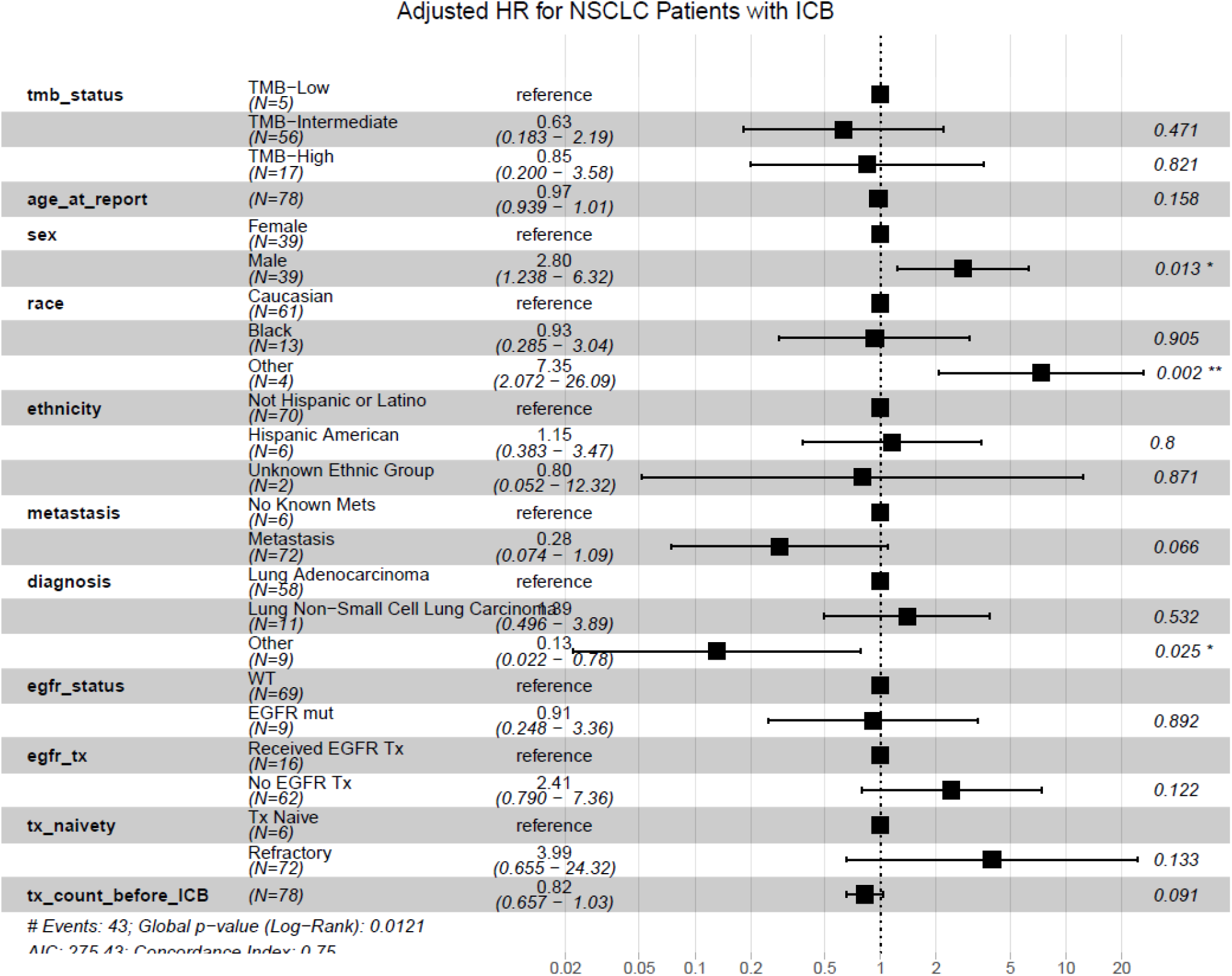
Hazard Ratios for TMB Status in NSCLC Patients with History of Immunotherapy Adjusted for Covariates.

## Discussion

We reported the TMB status and its association with OS in ICB treated NSCLC patients from a single institute, using TMB measures generated from routine clinical testing. A strength of our investigation is the use of TMB status measured by a routine clinical TGP. While whole exome sequencing and/or other measurements of TMB status could provide a more comprehensive technique for assessment of TMB within a sample, significant costs preclude routine use in clinical care at this time.^7^ Targeted sequencing assays such as FO, however, provide an acceptable cost and are approved by the FDA for clinical use, and thus have greater opportunity for widespread use. While previous studies have found an association between TMB status and survival outcomes for patients with ICB treatment^1,3,8^, there are still inconsistencies in the type of significant survival metric (OS, or progression-free survival) and also the thresholds that determine TMB-H vs TMB-L status. Single-system observational studies based on routine clinical care, such as the work in the present study, provide an opportunity to validate these associations with the possibility to control for important variables, such as treatment history and detailed diagnosis. They also allow us to complement clinical trial data by escaping some confounding factors that may be brought by inclusion criteria.

While our analysis showed no statistical significance of the TMB status with OS for ICB treated NSCLC patients, there are several limitations of our study. First, we did not consider different criteria of TMB status to categorize TMB high, intermediate, low patient subgroups. Our purpose of this study was to evaluate the prognostic utility of TMB status reported in the clinically available TGP report. Thus TMB status with different criteria and/or genomic profiling platforms including WEX should be evaluated. Secondly, we are limited to use of OS to evaluate the prognostic utility of TMB status in this study. The use of progression-free survival or Response Evaluation Criteria in Solid Tumors (RECIST) could provide more comprehensive evaluation of the prognostic utility of TMB to ICB response. Third, we did not investigate sub-stratified patient groups based on PD-L1 levels or ICB agent received, although PD-L1 levels may not be routinely available and studying differential outcomes across the ICB class as a whole provides a more real-world scenario. Lastly, our analysis is limited to OS from beginning of ICB treatment and not taking into account other treatments patients may have received during or after ICB treatments.

Nevertheless, our longitudinal-single institute analysis warrants caution on the use of TMB as a prognostic and/or predictive biomarker to guide ICB treatment in NSCLC and suggests that more and larger retrospective and prospective studies should be carried out to confirm the clinical utility of TMB as a prognostic and/predictive biomarker in the real-world setting.

## Data Availability

The datasets generated during and/or analysed during the current study are available from the corresponding author on reasonable request.

## Notes

### Competing Interest Statement

The authors have declared no competing interest.

### Funding Statement

No external funding was received.

